# An Interpretable Cost-Aware Framework for Mitigating Bias in Skin Lesion Classification Across Diverse Skin Tones

**DOI:** 10.1101/2024.12.11.24318858

**Authors:** Md. Mohit Hasan, Mahbuba Tasnime Suchi, Md. Hasibul Habib, Sumya Akter, David Eisenberg, Fahmida Zaman Achol, A.M. Tayeful Islam, Aaron Lotvola, Humayera Islam, Chelsey Hill, Jorge Fresneda Fernandez, Md. Golam Rabiul Alam, Tanmoy Sarkar Pias, Simon Bin Akter

## Abstract

Despite advances in dermatological AI, skin lesion predictions continue to exhibit significant bias, consistently exhibiting underperformance in brown and darker tones. This disparity stems largely from the lack of representation in commonly used datasets such as Fitzpatrick17k and Diverse Dermatology Images (DDI), which are heavily skewed toward lighter skin tones. Existing models, including those trained on balanced datasets like Skin Cancer Benign vs. Malignant, show steep drops in recall when evaluated across varying pigmentation. To address this inequity, the Cost-Aware EfficientNet (CAEN) model based on EfficientNet-B0, a widely adopted convolutional neural network, is proposed, incorporating attention mechanisms and custom cost functions to better capture underrepresented tones and reduce class imbalance. Incorporating an optimization-based augmentation strategy further enhances fairness by iteratively determining the optimal augmentation ratio for each skin tone, resulting in a more balanced training. CAEN achieved average-recall rates of 86% for non-neoplastic, 87% for benign, and 87% for malignant lesions in three datasets, with a performance gain of 16.75% over prior studies. The model also demonstrated robustness under artificial test conditions, including variations in brightness and contrast, with marked improvement for brown and darker skin tones. Interpretability analysis using Gradient-Weighted Class Activation Mapping (Grad-CAM) showed that baseline EfficientNet-B0 often focuses on irrelevant image regions in darker skin tones. CAEN corrected these issues by attending more accurately to areas relevant to the lesion, improving both transparency and equity. This approach marks a step toward fair and generalizable lesion prediction, addressing long-standing performance disparities across diverse pigmentations in dermatological AI.

**Impact Statement:** The Cost-Aware EfficientNet (CAEN) incorporates custom cost allocation, attention mechanisms, and skin-tone-specific augmentation, significantly enhancing recall rates and improving prediction accuracy across various skin types. The model also increases transparency using Grad-CAM, offering visual explanations for its decisions. Furthermore, CAEN demonstrates robustness and generalizability, even when faced with artificial image quality drops across multiple datasets. By prioritizing fairness, this study aims to provide a more reliable and equitable AI tool for dermatology, ensuring better healthcare outcomes for individuals of all skin tones.

---

Skin lesions [1], [2] affect individuals across all skin tones, but diagnosing them using artificial intelligence (AI) [3] prediction techniques [4], [5] can be difficult due to variations in pigmentation [6], [7], [8], [9]. The Fitzpatrick 17k [10] and Diverse Dermatology Images (DDI) [11] datasets from Stanford University provide a valuable resource for classifying skin diseases using AI [12], [13] approaches, as it covers a diverse range of skin tones [7], [9]. However, a major challenge in applying prediction techniques to these datasets is the imbalance among different skin tone classes [6], [7], [14], [15]. AI models are often found to exhibit bias toward certain skin tones, which can result in incorrect diagnoses, particularly for underrepresented skin tones, especially brown and darker shades [6], [7], [3], [14], [15], [16], [17].

Previous studies utilizing datasets such as Fitzpatrick 17k and DDI have applied AI models for skin disease classification, but these efforts have faced significant challenges, particularly when dealing with brown and darker skin tones [6], [7], [3], [8], [9], [14], [15], [16], [17]. This has raised important questions about the generalizability of these models across diverse populations, pointing to a critical gap in understanding their applicability and effectiveness across all skin tones. Many of these studies have primarily focused on improving performance in skin lesion detection [18] using accuracy as the key performance measure [6], [7]. However, this metric can hide how much better models perform on majority skin tones compared to minority skin tones, leading to biased results that favor lighter skin tones over brown and darker ones [19], [20].

The need for accurate diagnosis across all skin tones has made it essential to address the limitations of prior approaches [6], [7], [3], [14], [15], [16], [17]. The reliance on overall performance metrics like accuracy does not offer a reliable measure of fairness in class-wise predictions, especially when dealing with imbalanced datasets like Fitzpatrick 17k and DDI [20]. Therefore, it is essential to explore how imbalances can be more carefully addressed to improve fairness and to ensure more equitable predictions across all skin tones.

This study investigates the following research questions:

1. Do machine learning-based skin lesion classification models exhibit performance disparities across different skin tone groups, particularly between lighter and darker tones?
2. How does dataset imbalance, especially the dominance of lighter skin tone samples, affect the performance of skin lesion classification models?
3. How can predictive models be improved to better learn from underrepresented skin tone groups in the presence of limited and imbalanced data?
4. How can interpretability and performance consistency across diverse skin tone groups be enhanced in skin lesion classification models?

In this study, we have initially used the Skin Cancer: Malignant vs. Benign dataset provided by the International Skin Imaging Collaboration (ISIC) archive [21], which does not contain any skin tone variations or class imbalances. We have found that prediction models perform significantly well when there are no racial variations in the samples. However, when tested against datasets like the Fitzpatrick 17k and DDI datasets, which contain racial variations, the performance has significantly varied. To mitigate racial biases and class imbalances in the Fitzpatrick 17k and DDI datasets, we have explored various modeling approaches combined with an optimization-based augmentation technique to perform well in both scenarios, with and without racial biases. The primary contribution of this work is the proposed Cost-Aware EfficientNet (CAEN), which extends EfficientNet-B0 with spatial-channel attention and cost-sensitive optimization to improve lesion classification under class imbalance and skin tone variability. The framework is designed to enhance performance consistency across diverse skin tone groups while prioritizing lesion-specific feature learning. Additionally, the study emphasizes evaluating class-wise performance and its enhancement rather than relying solely on overall metrics. The CAEN model is based on the EfficientNet architecture and incorporates dynamic cost-sensitive learning (CSL) [22] and an attention mechanism [23], fine-tuned to predict skin lesions, including **i. non-neoplastic, ii. benign**, and **iii. malignant**, across six skin tones from light to dark. Additionally, we have tested the proposed model across different augmented samples from the test sets, varying brightness, contrast, and zoom, highlighting the importance of fairness in AI models [24] to ensure accurate predictions for all skin tones. This study also includes detailed interpretability [12], [13] of its predictions across different skin tones, ensuring transparency in how the model makes decisions for diverse skin tones using Gradient-Weighted Class Activation Mapping (Grad-CAM) [25].

The key contributions are summarized below.

1. The proposed attention-cost network improves sensitivity to underrepresented skin tones, such as dark and brown tones, thereby reducing performance disparities across groups.
2. Assigning higher misclassification costs to underrepresented tones improves learning from these minority groups by increasing their contribution during optimization.
3. The incorporation of attention mechanisms improves feature representation learning by emphasizing lesion-relevant regions across varying skin tone distributions.
4. The use of optimized sampling ratios generates synthetic data to improve class and skin tone balance, thereby enhancing model robustness.
5. Grad-CAM is applied to provide qualitative interpretability of model decisions across different skin tone groups by visualizing class-discriminative regions.

## A. Related Works And Their Limitations

AI-based skin lesion classification systems have demonstrated strong performance in controlled settings; however, multiple studies have reported significant performance disparities across different skin tone groups, particularly for brown and darker skin types [6], [7], [3], [8], [9], [14], [15], [16], [17], [26], [27]. These disparities are primarily attributed to dataset imbalance, underrepresentation of darker skin tones, and spurious correlations learned by deep learning models.

Existing fairness-aware approaches in dermatology can be broadly categorized into four major directions:

1. **Reweighting and cost-sensitive learning:** Several studies assign higher weights to minority classes or underrepresented demographic groups to reduce prediction bias [15], [17]. While effective in improving recall for minority groups, these methods often fail to capture complex feature-level disparities across skin tones.
2. **Data-level balancing and augmentation:** Some approaches attempt to mitigate bias by oversampling under-represented skin tones or using augmentation strategies to balance datasets [7], [15], [17]. However, such methods can introduce synthetic distribution shifts and do not explicitly address feature representation bias.
3. **Domain adaptation and distribution alignment:** Domain adaptation techniques aim to reduce inter-domain shifts between skin tone groups by aligning feature distributions across domains [14]. Despite their effectiveness, these methods often require explicit domain labels and can not generalize well under severe class imbalance.
4. **Fair representation learning and disentanglement:** Recent studies explore learning disentangled representations that separate identity-related attributes (e.g., skin tone) from disease-relevant features [15]. Although promising, these approaches introduce additional architectural complexity and often require strong supervision signals.

Despite these advancements, existing methods still face limitations in achieving consistent performance across all skin tone groups [6], [3], [9], [27]. For example, reported results on Fitzpatrick17k indicate substantial variability in recall across classes, with significantly lower performance observed for malignant cases in darker skin tones [6], [7], [15].

In contrast to prior work, the proposed CAEN integrates cost-sensitive learning with spatial-channel attention within a unified EfficientNet-B0 framework. Unlike reweighting-only approaches, CAEN combines sample-level cost adjustment with feature-level refinement, enabling the model to focus on lesion-specific structures rather than relying on dataset-induced biases [15], [17]. Furthermore, unlike disentanglement-based methods, CAEN does not require explicit attribute separation, reducing architectural complexity while maintaining robustness across heterogeneous skin tones [15].

Empirically, CAEN achieves a 16.75% improvement in overall performance compared to existing approaches [15], [17], [14], [7], [6], with consistent gains observed across both Fitzpatrick17k and DDI datasets. These improvements are particularly significant for minority skin tone groups, where prior methods exhibit substantial performance degradation. Additionally, CAEN demonstrates improved robustness under varying image conditions, highlighting its generalization capability across diverse dermatological datasets.

### This paper is structured as follows

The introduction discusses the challenges of skin lesion prediction in AI, particularly racial bias in predicting lesions on brown and dark skin tones. The methods section details the proposed CAEN model, which integrates CSL [22] and an attention mechanism to improve performance across diverse skin tones, even with limited data for dark and brown tones. It also explains the optimization-based sampling approach used to identify the ideal ratio for each skin tone of a particular class within a data set that balances the training set with synthetically created diverse skin tone data. The results section presents the CAEN model’s balanced performance on the Fitzpatrick 17k [10], DDI [11], and Skin Cancer: Malignant vs. Benign [21] datasets under varying brightness, contrast, and zoom test conditions. It also discusses the model’s interpretability, showing how the proposed model performs equally well on diverse skin tones. The discussion evaluates the model’s effectiveness in improving subgroup performance consistency across different skin tones while addressing class imbalance in dermatological datasets. Finally, the conclusion summarizes the contributions and implications for fairer AI in dermatology.

## I. MATERIALS AND METHODS

We have utilized the Fitzpatrick 17k and DDI datasets, which include three to six different skin tones ranging from light to dark, to predict skin lesions using AI-based approaches, as illustrated in **Fig 1**. The Fitzpatrick 17k dataset includes six skin tones (Type I to Type VI according to the Fitzpatrick scale), while the DDI dataset includes three tones (light, brown, and dark). To address the challenges associated with skin tone variability and class imbalance, we developed the proposed CAEN, which extends EfficientNet-B0 with attention-based feature refinement and CSL mechanisms. **Fig 1** presents a top-down view of the comprehensive workflow. First, the datasets have been preprocessed to remove noise and standardize image size and quality. Next, to mitigate class imbalance and improve model learning on underrepresented dark and brown skin tones, we applied an optimization-based sampling approach. This method involves generating synthetic data through augmentation (e.g., rotation, brightness adjustment, contrast adjustment, and zooming) to balance the representation of different skin tones. The ideal augmentation ratio for each skin tone from a particular class within a dataset has been determined through iterative testing to maximize balanced learning across all skin tones. The CAEN model has been trained and tested across these three datasets: Fitzpatrick 17k, DDI, and a balanced dataset (Skin Cancer: Malignant vs. Benign) that lacks skin tone variations. Then, the results have been compared with existing models that involve analyzing class-wise recall to identify improvements in predictive performance, particularly for brown and dark skin tones. To further assess the model’s adaptability and robustness, we evaluated performance under controlled test conditions, including brightness, contrast, and zoom variations. The CAEN model has demonstrated consistent and balanced performance across diverse skin tones and test conditions, highlighting its ability to generalize effectively. Furthermore, to ensure interpretability, we have applied Grad-CAM to visualize how the CAEN model prioritized features across different skin tones. This analysis has revealed that the model effectively focused on lesion-specific features rather than skin tone variations, demonstrating fairness and transparency in decision-making. This comprehensive evaluation has confirmed the robustness and reliability of the CAEN model for dermatological prediction across diverse real-world conditions.

**Fig. 1.**
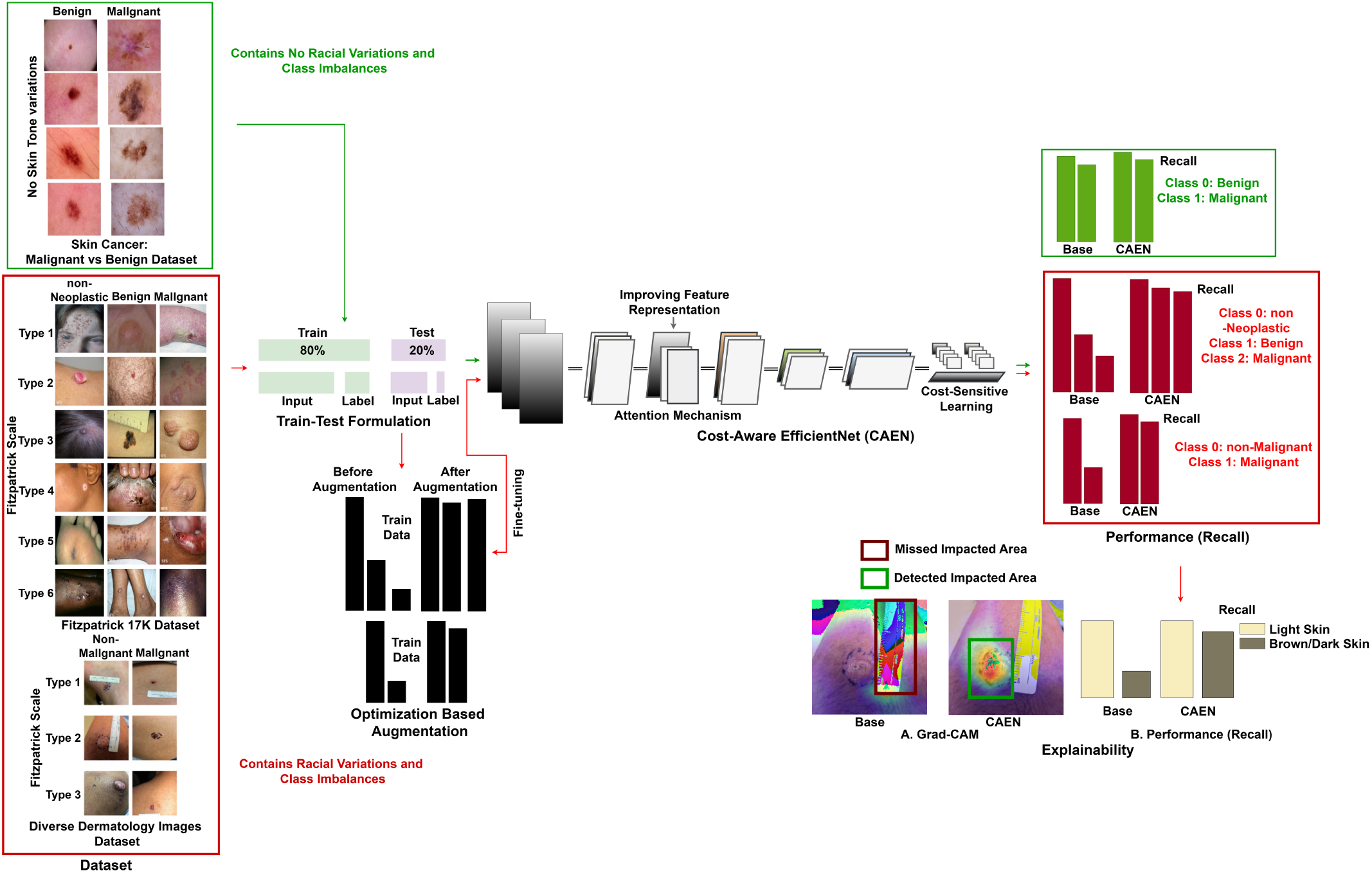
Comprehensive Workflow for Mitigating Racial Bias in Skin Lesion Detection. The process involves optimizing the augmentation ratio to achieve ideal performance for each skin tone from a class within a dataset, leveraging a Cost-Aware EfficientNet architecture to address class imbalances and racial biases, and conducting a thorough interpretability analysis.

### A. Data Description

The Fitzpatrick 17k [10] dataset comprises a diverse collection of skin disease images categorized by six distinct skin tones: **Type I (very fair), Type II (fair), Type III (medium), Type IV (olive), Type V (brown)**, and **Type VI (dark)**. In this study, we utilized a total of 16,012 images from this dataset [10] to predict skin lesions, specifically focusing on three categories: **(i) non-neoplastic, (ii) benign**, and **(iii) malignant** lesions. Additionally, we have employed the DDI dataset [11], which includes only three skin tones: **Type I (fair), Type II (brown)**, and **Type III (dark)**. From this dataset [11], we have used a total of 656 images for skin lesion prediction, specifically focusing on two categories: **(i) non-malignant**, and **(ii) malignant** lesions. However, it is important to note that the images for different skin lesions are heavily imbalanced in both datasets, with very limited samples available for brown and darker skin tones, posing challenges for accurate model training and evaluation depicted in **Table I**. Further, we have used a balanced dataset, Skin Cancer: Malignant vs. Benign [21], which contains no skin tone variations provided by the ISIC archive. We utilized a total of 3,297 images from this dataset to predict skin lesions, including **(i) benign** and **(ii) malignant**.

**TABLE I.** Training and testing distribution across datasets. The reported augmentation ratios are obtained from the Adaptive Sampling Optimization Strategy described in Section I-C. The final values correspond to convergence after *T* iterations under the stopping criteria defined in the same framework.

| Fitzpatrick Scale<br>(Skin Tone) | Train Set |  |  | Test Set |  |  |
| --- | --- | --- | --- | --- | --- | --- |
|  | Non-Neoplastic | Benign<br>(Non-Malignant) | Malignant | Non-Neoplastic | Benign<br>(Non-Malignant) | Malignant |
| Type I (very fair) | 1654 | 347 → 1884 | 365 → 927 | 396 | 97 | 88 |
| Type II (fair) | 2729 | 535 → 3108 | 589 → 2905 | 666 | 136 | 153 |
| Type III (medium) | 1901 | 377 → 2049 | 366 → 1391 | 476 | 98 | 90 |
| Type IV (olive) | 1692 | 295 → 1604 | 237 → 1264 | 421 | 72 | 64 |
| Type V (brown) | 973 | 137 → 744 | 120 → 636 | 254 | 22 | 27 |
| Type VI (dark) | 409 | 35 → 194 | 48 → 252 | 121 | 9 | 13 |
| Light | N/A | 112 → 319 | 37 → 325 | N/A | 47 | 12 |
| Brown | N/A | 115 → 331 | 54 → 477 | N/A | 52 | 20 |
| Dark | N/A | 112 → 316 | 29 → 256 | N/A | 47 | 19 |
| Light | N/A | 1440 | 1197 | N/A | 360 | 300 |

### B. Train/Test Formulation

All images are resized to 224 *×* 224 pixels and normalized to the range [0, 1] prior to training. The dataset is partitioned into training and testing sets using an 80:20 split ratio. To ensure unbiased evaluation and prevent data leakage, patient-level splitting is enforced such that images from the same patient are strictly confined to either the training or testing set, but not both. To improve data diversity and model generalization, standard geometric and photometric augmentation techniques are applied exclusively to the training set.

The applied augmentation operations are defined within the following clinically constrained ranges [28], [29]:

- **Rotation:** random rotations by multiples of 90° (0°, 90°, 180°, and 270°) and additional rotations within *±*15°
- **Horizontal and Vertical Flipping**
- **Brightness Adjustment:** scaling factor in [0.8, 1.2]
- **Contrast Adjustment:** scaling factor in [0.8, 1.2]
- **Translation:** up to 10% of the image dimensions
- **Scaling (Zooming):** range [0.8, 1.2]

These augmentation ranges are selected to preserve clinically relevant dermoscopic features based on clinically standard dermoscopic transformations reported in prior literature [28], [29], including lesion boundary structure, pigmentation distribution, and texture patterns, while avoiding unrealistic distortions that can introduce non-clinical artifacts. The same augmentation strategy and parameter ranges are applied uniformly across all skin tone groups and lesion classes to avoid introducing additional distributional bias between subgroups. The final augmentation ratios for each class and skin tone are determined using the Adaptive Sampling Optimization Strategy described in Section I-C.

### C. Adaptive Sampling Optimization Strategy

To address class imbalance across both lesion categories and Fitzpatrick skin tone subgroups, this study introduces an Adaptive Sampling Strategy that iteratively refines the augmentation ratio based on validation recall. Unlike gradient-based optimization, the proposed method follows a metric-guided discrete search process in which the sampling ratio is updated according to performance feedback from the validation set.

Let 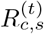 denote the sampling ratio for class *c* and skin tone *s* at iteration *t*. The initial sampling ratio is set uniformly as:

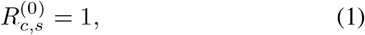

ensuring that all classes and skin tones contribute equally at the start of training.

The update rule is defined as:

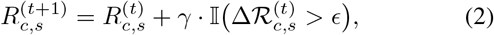

where *γ* is the growth factor, *I*(*·*) is an indicator function, and 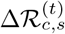 represents the change in validation recall:

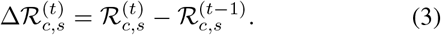

The parameter *ϵ* controls the minimum required improvement in recall for accepting an update. If 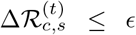, the sampling ratio is not increased, ensuring stability and preventing overfitting due to excessive augmentation.

The growth factor is set to *γ* = 0.5, which was empirically selected through preliminary experiments to balance convergence speed and augmentation stability.

#### Stopping Criteria

The iterative update process terminates when either (i) the maximum number of iterations *T* is reached or (ii) no improvement in recall is observed for *k* consecutive iterations:

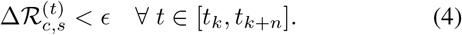

#### Convergence Behavior

Since the update rule operates in a discrete search space governed by validation performance, convergence is achieved when the sampling ratios stabilize and no further improvements in recall are observed across iterations.

#### Computational Complexity

The optimization introduces an additional overhead proportional to the number of iterations, resulting in a complexity of *O*(*T · N*), where *N* is the validation set size. However, this cost is incurred only during preprocessing and does not affect inference time.

Overall, this formulation transforms augmentation from a static preprocessing step into a performance-driven adaptive optimization process, ensuring better balance across both class labels and skin tone distributions.

### D. Proposed Cost-Aware EfficientNet

The proposed CAEN is an EfficientNet-B0-based architecture designed for skin lesion classification under conditions of class imbalance and demographic variability across skin tones. Rather than introducing a new backbone, CAEN extends EfficientNet-B0 with two key components: (i) a spatial-channel attention refinement module [23] for adaptive feature reweighting and (ii) a CSL [22] objective to mitigate class imbalance during optimization. The overall architecture is illustrated in **Fig 2**, where inherited EfficientNet components and newly introduced modules are explicitly distinguished.

**Fig. 2.**
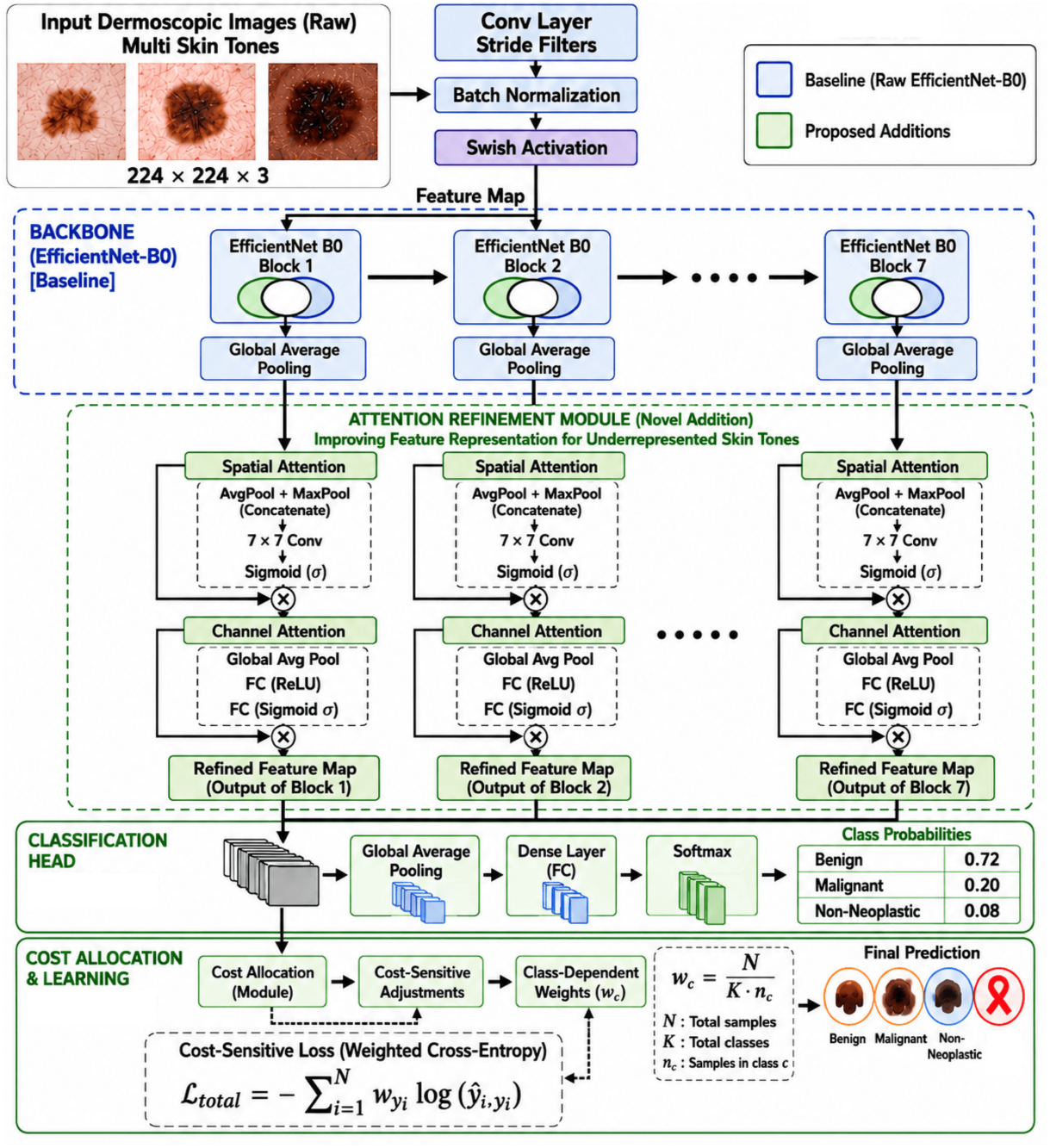
Architecture of the Cost-Aware EfficientNet (CAEN) model for Skin Lesion Classification. The CAEN model enhances the standard EfficientNet-B0 architecture by integrating cost-sensitive learning (CSL) and attention mechanisms to address biases associated with underrepresented brown and dark skin tones in skin lesion classification tasks. The architecture begins with a stem block, followed by a series of modified MBConv blocks that incorporate attention mechanisms to improve feature representation for underrepresented tones. CSL is applied during training to prioritize these underrepresented tones, ensuring balanced performance across diverse skin tones. The model concludes with a head block comprising global average pooling and a fully connected layer, leading to the final classification output. This design ensures equitable skin lesion predictions, contributing to fairer healthcare outcomes for diverse populations.

#### 1) Backbone Feature Extraction

Given an input dermoscopic image *I ∈* ℝ^224*×*224*×*3^, EfficientNet-B0 is used as a fixed backbone feature extractor to obtain high-level representations:

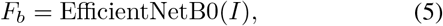

where *F*_*b*_ *∈* ℝ^*H×W ×C*^ denotes the extracted convolutional feature map. No modification is applied to the EfficientNet architecture; it serves solely as a pretrained feature encoder.

#### 2) Spatial Attention Module

To enhance lesion localization, a spatial attention mechanism is applied to emphasize discriminative regions within the feature map. The spatial attention map is computed as:

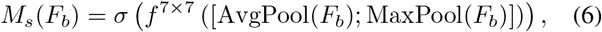

where AvgPool(*·*) and MaxPool(*·*) are channel-wise pooling operations, [*·*;*·*] denotes concatenation, *f* ^7*×*7^ is a convolutional layer, and *σ*(*·*) is the sigmoid function.

The refined spatial representation is given by:

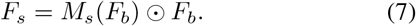

This operation adaptively enhances spatial regions that are statistically more informative for classification, such as lesion boundaries and irregular texture regions.

#### 3) Channel Attention Module

To model inter-channel dependencies and refine feature semantics, a channel attention mechanism is applied. First, global descriptors are computed as:

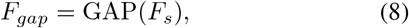

where GAP(*·*) denotes global average pooling. The channel attention weights are computed as:

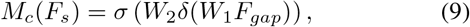

where *W*_1_ and *W*_2_ are learnable parameters and *δ*(*·*) is the ReLU activation function.

The final refined feature representation is:

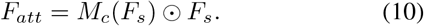

This formulation allows CAEN to suppress non-informative feature channels (often associated with background pigmentation variation) while enhancing lesion-relevant descriptors.

#### 4) Classification Head

The refined feature tensor *F*_*att*_ is passed through global average pooling and a fully connected layer followed by softmax activation:

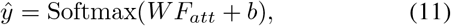

where *ŷ* denotes the predicted class probability distribution.

#### 5) Cost-Sensitive Learning Objective

To handle severe class imbalance, CAEN employs a weighted cross-entropy loss as the optimization objective. The total loss is defined as:

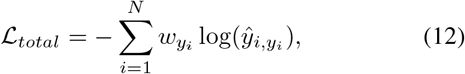

where *ŷ*_*i,yi*_ is the predicted probability for the true class *y*_*i*_, and *w*_*yi*_ is the class-dependent weight.

The class weights are defined as:

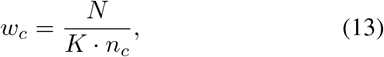

where *n*_*c*_ denotes the number of samples in class *c*.

This formulation increases the penalty for misclassification of minority classes, ensuring that the optimization process does not become biased toward majority classes. Unlike focal loss, which dynamically scales hard samples, the proposed formulation directly incorporates dataset-level class distribution into the loss function, making it more stable for medical datasets with extreme imbalance.

#### 6) Relationship to EfficientNet-B0 and Architectural Novelty

CAEN does not modify the internal convolutional structure of EfficientNet-B0. The architectural novelty lies in the insertion of two sequential refinement stages (spatial and channel attention) after feature extraction and the replacement of the standard cross-entropy objective with a cost-sensitive formulation.

Formally, EfficientNet-B0 acts as a fixed mapping function:

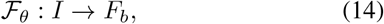

while CAEN extends this mapping as:

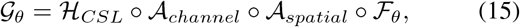

where *A*_*spatial*_ and *A*_*channel*_ denote the attention transformations and _*CSL*_ denotes cost-sensitive optimization.

#### 7) Bias Mitigation Rationale (Skin Tone Variability)

Although convolutional networks [6], [7], [3], [30], [31] naturally capture local spatial patterns, training on imbalanced dermatological datasets can introduce spurious correlations between class labels and skin-tone distributions. CAEN does not explicitly encode skin color information; instead, it reduces dependency on such correlations by strengthening attention toward lesion-specific morphological features, including asymmetry, border irregularity, texture variation, and intra-lesion color heterogeneity. Importantly, the proposed architecture does not claim explicit elimination of racial bias. Instead, it provides a mechanism that encourages representation learning to prioritize lesion characteristics over global background appearance. Any fairness improvement is validated empirically through subgroup evaluation across Fitzpatrick skin types rather than assumed from architectural design.

#### 8) Interpretability via Grad-CAM

The attention modules are internal feature reweighting mechanisms used during forward propagation, whereas Grad-CAM is employed post-hoc to visualize class-discriminative regions. Specifically, Grad-CAM provides gradient-based localization of influential spatial regions in the final convolutional layer, allowing qualitative verification that the learned attention aligns with lesion regions. Thus, attention guides feature learning, while Grad-CAM provides interpretability of the trained model behavior.

#### 9) Training Setup and Model Complexity

All experiments were conducted using TensorFlow/Keras on Google Colab with a Tesla T4 GPU. On the Fitzpatrick17K dataset, EfficientNet-B0 contains 5,368,486 parameters, whereas CAEN contains 5,574,646 parameters (an increase of 206,160 parameters). On the DDI dataset, EfficientNet-B0 contains 4,052,133 parameters, while CAEN contains 5,573,621 parameters (an increase of 1,521,488 parameters). Despite this moderate increase in complexity, CAEN consistently produces a higher number of correct predictions across test sets, demonstrating that performance gains are attributable to the proposed attention refinement and cost-sensitive optimization rather than parameter scaling alone.

All models, including the proposed CAEN and baseline architectures, are trained using the Adam optimizer with an initial learning rate of 1 *×* 10^*−*4^. The batch size is set to 32, and training is performed for a maximum of 100 epochs. A cosine annealing learning rate scheduler is employed to stabilize convergence by gradually reducing the learning rate during training. To prevent overfitting, early stopping is applied based on validation loss with a patience of 10 epochs. The model with the best validation performance is retained for final evaluation.

## II. RESULTS AND ANALYSES

The performance of various models, including the proposed CAEN, has been evaluated using three distinct dermatological datasets: the Skin Cancer: Malignant vs Benign dataset, the Fitzpatrick 17k dataset, and the DDI dataset. We have evaluated the models on the Skin Cancer dataset, which lacks class imbalances and skin tone variations. Subsequently, we have tested the models on the Fitzpatrick 17k and DDI datasets, which are characterized by class imbalances and racial diversity in skin tones. These evaluations have been conducted to assess the models’ performance across varied real-world conditions. To estimate uncertainty, 95% confidence intervals are computed using a non-parametric bootstrap procedure with 1,000 resampling iterations. This approach is selected due to the presence of class imbalance and small subgroup sample sizes, which can violate normality assumptions required for parametric confidence intervals. For small subsets such as Fitzpatrick Type VI malignant cases, bootstrap-based confidence intervals are preferred over Gaussian approximations to provide more robust and distribution-free uncertainty estimates. To ensure statistical reliability, all reported performance metrics for the CAEN model are computed over 5 independent runs with different random seeds. The mean and standard deviation are reported to capture performance variability under stochastic training conditions. This evaluation is specifically conducted for CAEN to assess its robustness under artificial image conditions. Additionally, an ablation study has been conducted on both the Fitzpatrick17K and DDI datasets to evaluate how each component of the proposed CAEN contributes to overall performance, fairness, and robustness. Next, we have performed explainability analysis to further demonstrate the reliability of the models’ performance and their generalizability to real-world scenarios. This analysis has provided valuable insights into the models’ decision-making processes, highlighting their robustness in handling diverse and imbalanced dermatological datasets.

### A. Experimental Results

A class-wise recall comparison of various models using the Skin Cancer: Malignant vs Benign dataset has been presented in **Table II**. Most models, including the proposed CAEN, have performed similarly well, as this dataset has not included variations in skin tones or class imbalances. The confidence scores in **Table II** have also shown little variation, which has demonstrated competitive performance and strong generalizability. However, it is necessary to explore how these models perform when there are class imbalances and racial variations in the data, as this would be more accurate to real-world scenarios.

**TABLE II.** Class-wise recall comparison of the proposed model with existing approaches using the Skin Cancer: Malignant vs Benign dataset. This dataset does not include any variations in skin tones or class imbalances. Balanced Recall is computed as the average of Recall-Benign and Recall-Malignant, where higher values indicate better and more balanced class-wise detection performance. The 95% confidence interval is provided in parentheses.

| Model | Recall-Benign | Recall-Malignant | Accuracy (%) | Balanced Recall |
| --- | --- | --- | --- | --- |
| Contrastive Language-Image Pre-training | 0.83 (± 0.04) | 0.49 (± 0.05) | 67.43 | 0.66 |
| Inception ResNet V2 | 0.64 (± 0.05) | 0.76 (± 0.05) | 69.39 | 0.70 |
| Inception V3 | 0.78 (± 0.04) | 0.74 (± 0.05) | 75.76 | 0.76 |
| NASNetLarge | 0.81 (± 0.04) | 0.72 (± 0.05) | 76.97 | 0.77 |
| Xception | 0.84 (± 0.04) | 0.82 (± 0.04) | 82.73 | 0.83 |
| MobileNet V2 | 0.91 (± 0.03) | 0.75 (± 0.05) | 83.49 | 0.83 |
| VGG 19 | 0.86 (± 0.04) | 0.84 (± 0.04) | 85.00 | 0.85 |
| EfficientNet-B0 | 0.89 (± 0.03) | 0.86 (± 0.04) | 87.43 | 0.88 |
| ResNet 50 | **0.91 (± 0.03)** | 0.85 (± 0.04) | **89.09** | 0.88 |
| CAEN | 0.86 (± 0.04) | **0.91 (± 0.03)** | 87.88 | **0.89** |

A class-wise recall comparison of various models using the Fitzpatrick 17k dataset has been presented in **Table III**. Due to class imbalances and racial variations in skin tones, the performance of models that have performed well in **Table II** has significantly dropped, especially for brown and darker skin tones. However, the proposed CAEN model has demonstrated significantly balanced performance across all classes and skin types, with particularly strong results for brown and darker tones. The confidence scores reported in **Table III** provide an indication of the prediction certainty of the proposed CAEN model across diverse and imbalanced dataset conditions. The relatively stable confidence distributions suggest consistent model behavior across different samples and skin tone groups, complementing the quantitative performance results.

**TABLE III.** Class-wise recall comparison of the proposed model with existing approaches using the Fitzpatrick 17k dataset. The model is trained on all skin tones jointly. The test sets are categorized into light (I-II), brown (III-IV), and dark (V-VI) skin tones, with class-wise recall reported for non-neoplastic, benign, and malignant classes. Balanced Recall is defined as the average of class-wise recalls across the three diagnostic classes (non-neoplastic, benign, and malignant), representing fairness in terms of equal sensitivity across all classes; higher balanced recall indicates more balanced multi-class performance. The 95% confidence interval is included in parentheses.

| Model | Recall-Non-Neoplastic | Recall-Benign | Recall-Malignant | Accuracy (%) | Balanced Recall |
| --- | --- | --- | --- | --- | --- |
| Inception ResNet V2 | 0.01 (± 0.00) | 0.01 (± 0.00) | **0.92 (± 0.01)** | 16.11 | 0.31 |
| Contrastive Language-Image Pre-training | 0.14 (± 0.01) | 0.41 (± 0.02) | 0.07 (± 0.01) | 66.32 | 0.21 |
| NASNetLarge | 0.74 (± 0.02) | 0.20 (± 0.01) | 0.36 (± 0.02) | 58.29 | 0.43 |
| Inception V3 | 0.69 (± 0.02) | 0.27 (± 0.02) | 0.38 (± 0.02) | 54.43 | 0.45 |
| Xception | 0.69 (± 0.02) | 0.35 (± 0.02) | 0.37 (± 0.02) | 56.03 | 0.47 |
| MobileNet V2 | 0.57 (± 0.02) | 0.37 (± 0.02) | 0.53 (± 0.02) | 64.33 | 0.49 |
| VGG 19 | 0.83 (± 0.01) | 0.50 (± 0.02) | 0.62 (± 0.02) | 76.03 | 0.65 |
| EfficientNet-B0 | 0.84 (± 0.01) | 0.50 (± 0.02) | 0.70 (± 0.02) | 77.93 | 0.68 |
| ResNet 50 | **0.88 (± 0.01)** | 0.41 (± 0.02) | 0.70 (± 0.02) | 81.15 | 0.66 |
| CAEN (Light) | 0.75 (± 0.02) | 0.86 (± 0.02) | 0.75 (± 0.02) | 76.43 | 0.79 |
| CAEN (Brown) | 0.85 (± 0.03) | 0.88 (± 0.03) | 0.84 (± 0.03) | 84.53 | **0.86** |
| CAEN (Dark) | 0.74 (± 0.07) | 0.88 (± 0.05) | 0.86 (± 0.05) | **84.98** | 0.83 |
| CAEN (Overall) | 0.79 (± 0.01) | **0.88 (± 0.01)** | 0.81 (± 0.01) | 81.98 | 0.83 |

A class-wise recall comparison using the DDI dataset has demonstrated how the proposed CAEN model has achieved consistently strong performance across all skin tones, particularly for malignant cases represented in **Table IV**. Unlike CAEN, other models have struggled to handle variations in skin tones and class imbalances effectively, further validating CAEN’s robustness and adaptability. The results, including the confidence intervals reported in **Table IV**, reflect the stability of the proposed CAEN model’s performance across different skin tone groups under repeated evaluations. The relatively narrow confidence intervals suggest limited variability in performance estimates across skin tone-based subsets.

**TABLE IV.** Class-wise recall comparison of the proposed model with existing approaches using the Diverse Dermatology Images dataset. The model is trained on all skin tones jointly, and evaluated across light, brown, and dark skin tone subsets. Class-wise recall is reported for non-malignant and malignant classes, along with overall accuracy. Balanced Recall is defined as the mean of class-wise recalls (non-malignant and malignant), representing fairness in terms of equal sensitivity across both classes; higher balanced recall indicates more balanced performance between classes. The 95% confidence interval is included in parentheses.

| Model | Recall<br>Non-Malignant | Recall-Malignant | Accuracy<br>(%) | Balanced Recall |
| --- | --- | --- | --- | --- |
| Contrastive Language-Image Pre-training | 0.78 (± 0.06) | 0.18 (± 0.05) | 61.95 | 0.48 |
| Inception V3 | 0.68 (± 0.07) | 0.36 (± 0.07) | 59.39 | 0.52 |
| MobileNet V2 | 0.74 (± 0.06) | 0.43 (± 0.07) | 65.98 | 0.59 |
| VGG 19 | 0.73 (± 0.06) | 0.45 (± 0.07) | 65.97 | 0.59 |
| EfficientNet-B0 | 0.76 (± 0.06) | 0.45 (± 0.07) | 68.00 | 0.61 |
| NASNetLarge | 0.81 (± 0.05) | 0.30 (± 0.06) | 67.50 | 0.56 |
| Xception | 0.84 (± 0.05) | 0.20 (± 0.06) | 67.01 | 0.52 |
| Inception ResNet V2 | 0.89 (± 0.04) | 0.18 (± 0.05) | 70.04 | 0.54 |
| ResNet 50 | 0.86 (± 0.05) | 0.49 (± 0.07) | 76.16 | 0.68 |
| **CAEN (Light)** | 0.92 (± 0.07) | **1.00 (± 0.00)** | **93.28** | **0.96** |
| **CAEN (Brown)** | 0.90 (± 0.07) | 0.95 (± 0.05) | 91.67 | 0.93 |
| **CAEN (Dark)** | **0.98 (± 0.03)** | 0.79 (± 0.10) | 92.43 | 0.89 |
| **CAEN (Overall)** | 0.93 (± 0.04) | 0.91 (± 0.04) | 92.46 | 0.92 |

The confusion matrices (CM) across the DDI dataset have shown that the CAEN model has achieved strong performance across all classes, particularly in handling variations in skin tones and class imbalances represented in **Fig 3**. Compared to other models, the CAEN model has demonstrated superior classification performance, highlighting its robustness and effectiveness in real-world dermatological image analysis.

**Fig. 3.**
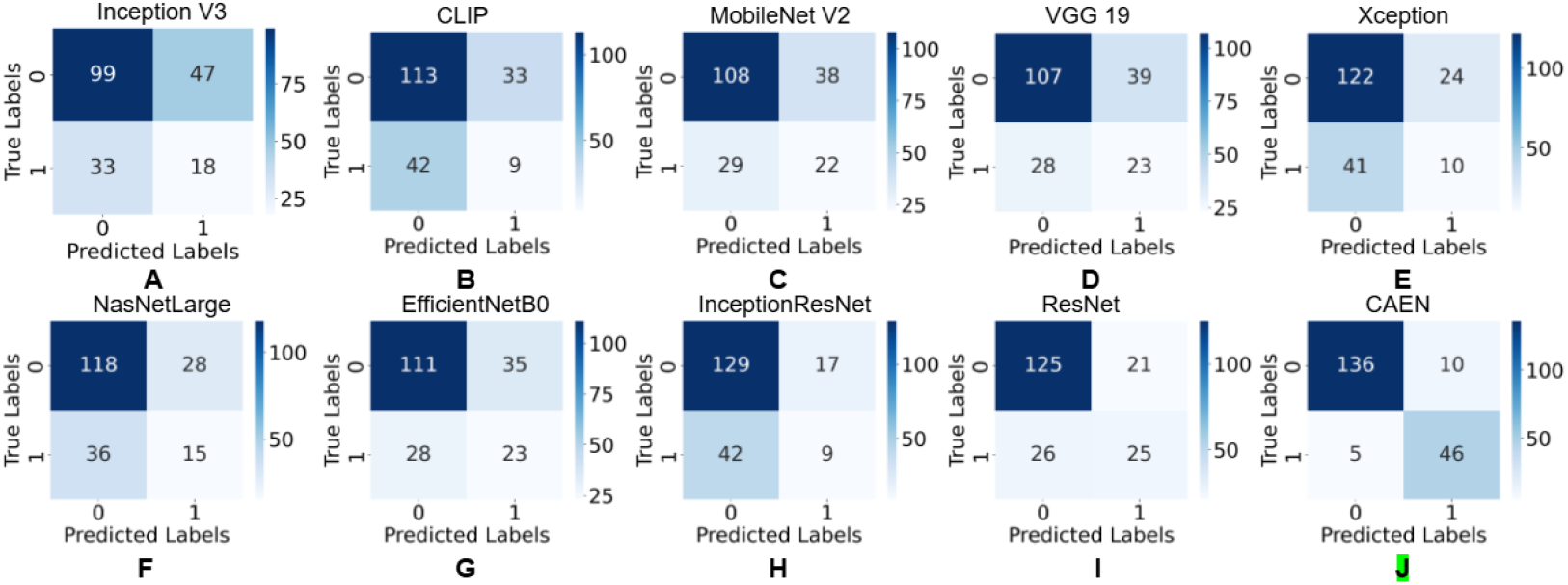
Confusion matrices (CM) across the Diverse Dermatology Images (DDI) dataset. (A) CM for Inception Version 3. (B) CM for Contrastive Language-Image Pre-training (CLIP). (C) CM for Mobile Neural Network (MobileNet) Version 2. (D) CM for Visual Geometry Group (VGG) 19. (E) CM for Xception. (F) CM for Neural Architecture Search Network - Large (NasNetLarge). (G) CM for Efficient Neural Network - Baseline Version 0 (EfficientNetB0). (H) CM for Inception-Residual Network (InceptionResNet). (I) CM for Residual Network (ResNet). (J) CM for Cost-Aware Efficient Neural Network (CAEN).

In **Table V**, for each model, recall is computed separately for Light, Brown, and Dark skin tone groups. In Fitzpatrick17K, recall is first computed for the three classes (non-neoplastic, benign, and malignant) and then averaged to obtain a single performance score for each skin tone group. In DDI, recall is computed for the two classes (non-malignant and malignant) and averaged to obtain the corresponding skin-tone-level score. Using these aggregated values, the Fairness Gap is calculated as the difference between the maximum and minimum recall across skin tone groups. Worst-Group Recall represents the lowest recall achieved among all groups, while Worst Group identifies the corresponding skin tone group. Bias Pattern qualitatively summarizes whether model performance is skewed toward a particular skin tone group or remains relatively balanced across groups. These metrics are computed independently for each dataset to evaluate subgroup performance consistency and robustness across demographic groups. All metrics are computed using the held-out test sets and are reported separately for each dataset to avoid cross-dataset aggregation effects.

**TABLE V.** Cross-dataset fairness evaluation across Fitzpatrick17K and DDI datasets. Fairness is quantified using (i) *Fairness Gap*, defined as the difference between maximum and minimum recall across skin tone groups (Light, Brown, Dark), where lower values indicate more consistent performance; (ii) *Worst-Group Recall*, representing the minimum recall achieved among all skin tone groups; and (iii) *Bias Pattern*, which qualitatively summarizes whether model performance is skewed toward a particular skin-tone group or remains relatively balanced across groups. Models with lower Fairness Gap and higher Worst-Group Recall are considered to exhibit more consistent performance across demographic groups.

| Model | Fitzpatrick17K |  |  | DDI |  |  | Bias Pattern |
| --- | --- | --- | --- | --- | --- | --- | --- |
|  | Fairness Gap | Worst Recall | Worst Group | Fairness Gap | Worst Recall | Worst Group |  |
| Inception ResNet V2 | 1.00 | 0.00 | Poor | 1.00 | 0.00 | Poor | Failed |
| EfficientNet-B0 | 0.91 | 0.00 | Brown/Dark | 0.73 | 0.17 | Light | Light |
| NASNetLarge | 0.61 | 0.13 | Brown | 0.71 | 0.17 | Light | Inconsistent |
| ResNet50 | 0.63 | 0.27 | Dark | 0.55 | 0.35 | Brown | Moderate |
| Xception | 0.44 | 0.24 | Dark | 0.77 | 0.10 | Brown | Unstable |
| MobileNet V2 | 0.46 | 0.33 | Brown | 0.44 | 0.37 | Dark | Mild Dark |
| VGG19 | 0.48 | 0.39 | Light | 0.38 | 0.25 | Light | Brown/Dark |
| Contrastive Language-Image Pre-training | 0.23 | 0.01 | Dark | 0.27 | 0.16 | Dark | Weak |
| Inception V3 | 0.52 | 0.19 | Light | 0.20 | 0.22 | Light | Mixed |
| CAEN | **0.14** | **0.74** | Dark | **0.19** | **0.79** | Dark | **Balanced** |

This analysis indicates that the proposed CAEN model achieves more consistent subgroup performance than the evaluated baseline methods across both Fitzpatrick17K and DDI datasets. While several conventional architectures, including Inception ResNet V2, EfficientNet-B0, ResNet50, and MobileNet V2, exhibit larger fairness gaps and lower worst-group recall values, CAEN consistently achieves the lowest fairness gap and the highest worst-group recall across both datasets. These results suggest reduced performance disparity across Light, Brown, and Dark skin tone groups while maintaining strong predictive performance. Furthermore, CAEN exhibits a more balanced distribution of subgroup performance, indicating improved robustness to demographic variability compared with the evaluated baseline approaches.

The **Table VI** presents the robustness analysis and ablation study results on the Fitzpatrick17K and DDI datasets. To evaluate robustness under real-world perturbations, standard deviations are reported for artificially altered image qualities, including variations in brightness, contrast, and zoom. On the Fitzpatrick17K dataset, the proposed model exhibits low variability with standard deviations of *±*0.04 for non-neoplastic cases, *±*0.01 for benign cases, and *±*0.04 for malignant cases. Similarly, on the DDI dataset, the model maintains stable performance with standard deviations of *±*0.03 for non-malignant and *±*0.04 for malignant cases, indicating consistent behavior under input perturbations.

**TABLE VI.** Robustness and ablation analysis of the proposed caen model across the Fitzpatrick17K and DDI datasets. Standard deviations (*±*SD) are computed from recall values obtained under artificial image quality variations including brightness, contrast, and zoom adjustments. The ablation study evaluates the contribution of cost-sensitive learning (CSL) and the attention mechanism to overall recall performance.

| Dataset | Class | Robustness (±SD) | Ablation Study (Recall) | | | |
| --- | --- | --- | --- | --- | --- | --- |
|  |  |  | EfficientNet-B0 | + CSL | + Attention | Final CAEN |
| Fitzpatrick17K | Non-Neoplastic | $\pm 0.04$ | 0.68 | 0.73 | 0.75 | 0.83 |
| | Benign | $\pm 0.01$ | | | | |
| | Malignant | $\pm 0.04$ | | | | |
| DDI | Non-Malignant | $\pm 0.03$ | 0.60 | 0.77 | 0.83 | 0.92 |
| | Malignant | $\pm 0.04$ | | | | |

In addition, the ablation study highlights the incremental contribution of each component of the proposed CAEN framework. On Fitzpatrick17K, the baseline EfficientNet-B0 achieves a recall of 0.68. The introduction of cost-sensitive learning improves recall to 0.73, while the inclusion of the attention mechanism alone further increases it to 0.75. When both components are integrated, the full CAEN model achieves a final recall of 0.83, demonstrating their complementary effects. A similar trend is observed on the DDI dataset, where the baseline model achieves a recall of 0.60. Cost-sensitive learning increases this to 0.77, attention mechanism improves it to 0.83, and the combined CAEN model achieves a final recall of 0.92. These results clearly demonstrate that each proposed component contributes significantly to improving predictive performance and robustness across both datasets.

### B. Explainability

The Grad-CAM visualizations in **Fig 4** provide qualitative insights into the regions influencing model predictions across different datasets. The affected lesion regions are generally highlighted with higher activation values, while lower activation areas correspond to less influential regions in the decision-making process.

**Fig. 4.**
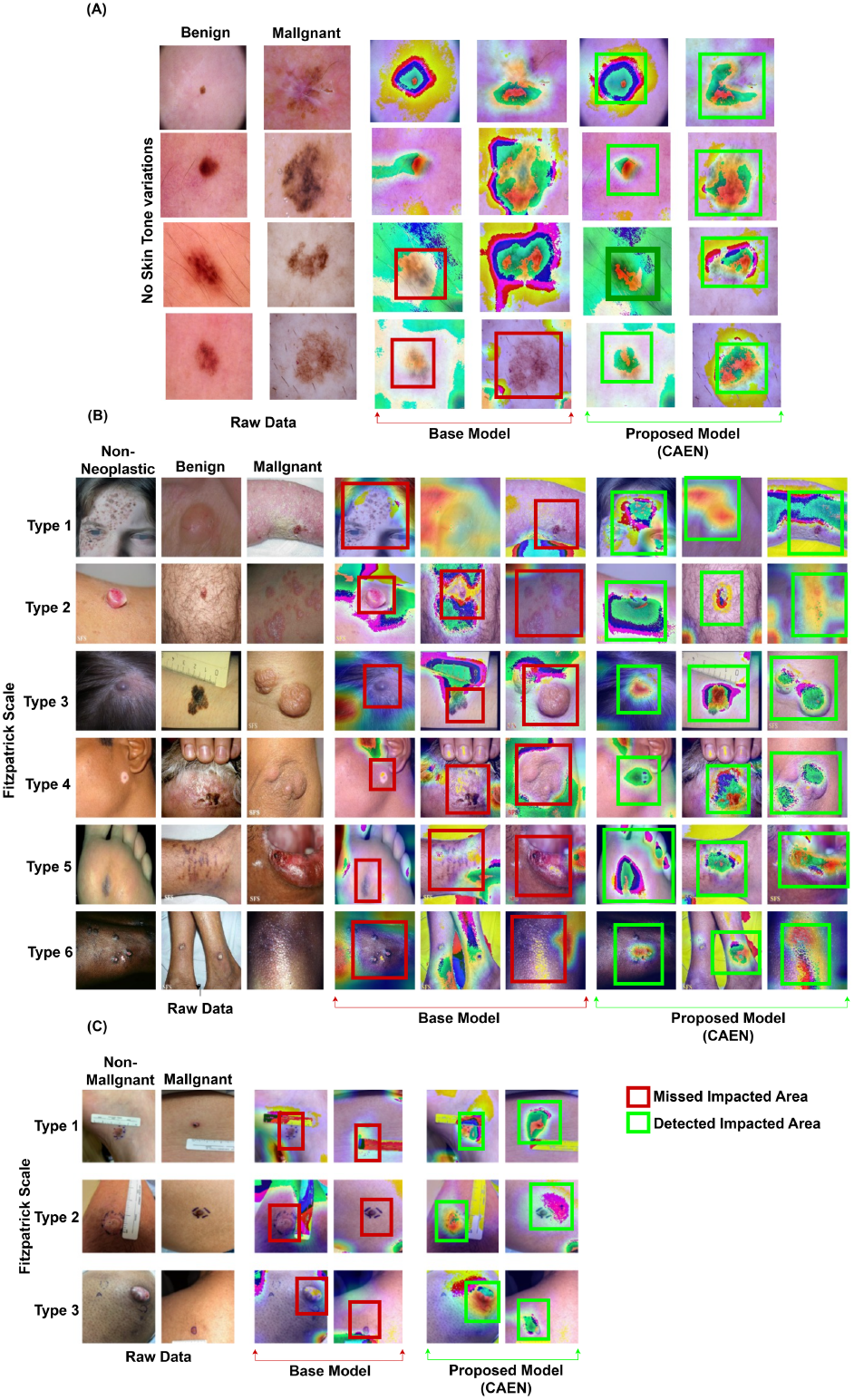
Explainability analysis of the CAEN model compared with the base model (EfficientNet-B0) using Gradient-weighted Class Activation Mapping (Grad-CAM). (A) Explainability across the skin cancer: malignant vs. benign dataset. (B) Explainability across the Fitzpatrick 17k dataset. (C) Explainability across the Diverse Dermatology Images (DDI) dataset. This analysis highlights how the base model has struggled to identify affected areas across different skin tones, whereas the proposed CAEN model has effectively addressed this limitation. Green highlights the most important regions, while orange and yellow indicate the critical areas.

Across the Fitzpatrick17k and DDI datasets, the base EfficientNet model shows less consistent focus on lesion-relevant regions, particularly in cases involving darker skin tones, where attention can extend to surrounding or non-lesion areas. In contrast, the proposed CAEN model demonstrates more consistent localization of lesion-relevant regions across different skin tone groups.

However, it is important to note that Grad-CAM provides qualitative interpretability and does not directly measure model fairness or classification correctness. It is used here to complement quantitative results reported in **Table III, Table IV**, and **Table V**.

Overall, these visualizations suggest that CAEN improves the consistency of feature localization across diverse skin tone groups compared to the baseline model, supporting the quantitative findings of improved subgroup performance stability.

## III. DISCUSSION

This study has highlighted the significant performance disparities that exist in dermatological prediction models, particularly in the context of skin tone variations and class imbalances. We have observed that many existing models, such as InceptionV3, ResNet50, and EfficientNet, show a notable decline in performance when applied to datasets with diverse skin tones, especially brown and darker tones represented in **Table III, Table IV**, and **Table V**. This performance variation indicates that models trained on imbalanced dermatological datasets are sensitive to demographic distribution shifts, which can adversely affect minority skin tone groups.

Although optimization-based augmentation was applied to address class and skin tone imbalance, as reflected in the improved data distributions in **Table II, Table III, Table IV**, and **Table V**, baseline models still exhibit reduced performance on brown and darker skin tones. This suggests that data-level balancing alone is insufficient to fully eliminate subgroup performance disparities in dermatological classification tasks. The observed discrepancy is particularly evident in models with lower recall for minority skin tone groups, further emphasizing the importance of evaluating performance beyond overall accuracy as discussed in [20].

The accuracies of several models reported in **Table III** and **Table IV** appear relatively high; however, the corresponding recall scores remain significantly lower for underrepresented groups. This indicates that accuracy alone is not a reliable metric for evaluating performance in imbalanced medical datasets, and subgroup-level evaluation is necessary for a more comprehensive assessment [20].

To better understand model behavior, explainable AI techniques such as Grad-CAM have been applied. As shown in **Fig 4**, the baseline EfficientNet model often fails to consistently focus on lesion-relevant regions across different skin tones, particularly in brown and darker skin images. Instead, attention is sometimes diverted toward irrelevant background regions, contributing to misclassification. In contrast, the proposed CAEN model, which integrates CSL and an attention mechanism, demonstrates improved localization of lesion-relevant features across skin tone variations.

The Grad-CAM visualizations in **Fig 4** suggest that CAEN produces more consistent activation patterns on clinically meaningful regions across different skin tones. However, these visualizations provide qualitative interpretability and should not be interpreted as direct quantitative evidence of fairness or bias removal.

Overall, the results in **Table II, Table III, Table IV**, and **Table V** demonstrate that CAEN improves performance consistency across diverse skin tone groups compared to baseline models. The integration of cost-sensitive learning and attention-based feature refinement contributes to improved subgroup-level robustness and reduced performance disparity, while maintaining competitive overall classification performance.

Despite these improvements, certain limitations remain. The availability of malignant samples in darker skin tones is still limited, which can affect generalization performance. Additionally, external validation on independent clinical datasets was not performed. Future work should focus on expanding dataset diversity and conducting cross-institutional validation to further assess robustness and fairness in real-world clinical settings.

## IV. CONCLUSION

This study has highlighted the performance disparities present in dermatological prediction models, where AI systems often exhibit varying performance across different skin tone groups, with reduced recall observed for brown and darker skin tones in imbalanced datasets. These observations indicate that dataset imbalance and uneven representation across skin tones can significantly influence model generalization, particularly in underrepresented demographic groups.

The proposed approach addresses these challenges by integrating cost-sensitive learning and attention-based feature refinement, enabling improved subgroup-level performance consistency across diverse skin tones. This leads to more balanced predictions compared to baseline models, particularly in scenarios involving class imbalance and demographic variability. The results demonstrate improved robustness and more consistent performance across skin tone groups, contributing to more reliable dermatological image analysis in real-world settings. The proposed framework also enhances interpretability by providing clearer insights into model decision-making, which is essential for clinical trust and adoption. Overall, the approach contributes to ongoing efforts in equitable healthcare, supporting improved diagnostic reliability for underrepresented populations. Its ability to adapt across varying skin tone distributions can assist healthcare [32], [33], [34] professionals in reducing misclassification risks in diverse clinical environments. Furthermore, the model’s explainability supports better understanding of predictions, which is important for informed medical decision-making.

However, a key limitation of this study is the limited availability of malignant cases in darker skin tones. Initially, StyleGAN v3 [35], [36] was explored for synthetic data generation. Although it aimed to preserve lesion characteristics and skin tone distribution, experimental results on the Fitzpatrick17k dataset indicated that it was not consistently able to generate high-fidelity samples in most cases [37]. The overall quality and realism of the generated images were limited, largely due to constraints in the available training data [38]. Consequently, recall for malignant cases in darker skin tones remained relatively low, ranging between 0.42 and 0.48, while the Fréchet Inception Distance (FID) [39] values ranged between 63 and 65, indicating limited synthetic image quality. Therefore, this approach did not significantly improve performance for malignant cases in darker skin tones [40].

To address this limitation, an optimization-based augmentation strategy tailored to each skin tone within specific classes was adopted to improve data balance and model robustness. Future work will focus on addressing data scarcity in underrepresented skin tone groups by exploring more advanced generative models capable of producing high-quality synthetic data even under limited training conditions. This will help improve representation of malignant cases in darker skin tones and further enhance model generalization.

Overall, despite challenges in data availability for malignant cases in darker skin tones, this study demonstrates improved subgroup performance consistency and reduced disparity across skin tone groups in dermatological AI models. The results suggest the effectiveness of the proposed framework in improving balanced prediction behavior, while also highlighting important directions for future fairness-aware medical imaging research.

## Data Availability

Studies of publicly available human data
Link: https://skincon-dataset.github.io/

